# Designing research studies in writer’s cramp dystonia: an analysis of automated writing measures

**DOI:** 10.1101/2021.03.02.21252036

**Authors:** Noreen Bukhari-Parlakturk, Michael Lutz, Alec McConnell, Hussein Al-Khalidi, Joyce En-Hua Wang, Burton Scott, Pichet Termsarasab, Lawrence Appelbaum, Nicole Calakos

## Abstract

**Background:** Writer’s cramp (WC) dystonia presents with abnormal postures during the task of writing and is an ideal dystonia subtype to study disease mechanisms for all forms of focal dystonia. Development of novel therapies is contingent on identifying sensitive and specific measures that can relate to the clinical syndrome and achieve a realistic sample size to power clinical research study for a rare disease. Although there have been prior studies employing automated measures of writing kinematics, it remains unclear which measures can distinguish WC subjects with high sensitivity and specificity and how these measures relate to clinician rating scales and patient-reported disability. The goal of this study was to: 1-identify automated writing measures that distinguish WC from healthy subjects, 2-measure sensitivity and specificity of these automated measures as well as their association with established dystonia rating scales, and 3-determine the sample size needed for each automated measure to power a clinical research study.

**Methods:** 21 WC dystonia and 22 healthy subjects performed a sentence-copying assessment on a digital tablet in a kinematic software and hand recognition software. The sensitivity and specificity of automated measures was calculated using a logistic regression model. Measures were then correlated with examiner and patient rating scales. Power analysis was performed for 2 clinical research designs using these automated measures.

**Results:** Of the 23 automated writing measures analyzed, only 3 measures showed promise for use in a clinical research study. The automated measures of writing legibility, duration, and peak acceleration were able to distinguish WC from healthy controls with high sensitivity and specificity, correlated with examiner-rated dystonia sub-score measures and demonstrated relatively smaller sample sizes suitable for research studies in a rare disease population.

**Discussion:** We identified novel automated writing outcome measures for use in clinical research studies of WC subjects which capture key aspects of the clinical disease and can serve as important readout of dystonia disease mechanism as well as future disease interventions.

## INTRODUCTION

Dystonia is an involuntary movement disorder characterized by sustained or intermittent muscle contractions that lead to abnormal postures (Albanese et al. 2013; Goldman 2015; Jedynak, Tranchant, and Zegers de Beyl 2001). Dystonia can be generalized in the whole body or focal to a single body part. Of the two, focal dystonias are more common (Ortiz et al. 2018; Nutt et al. 1988; Warner et al. 2000). Task specific focal dystonias are a fascinating subset that occur during a specific motor task. Writer’s cramp (WC) is an example of a task-specific focal hand dystonia that occurs during the select motor task of writing (Hallett 2006). There are no disease-modifying therapies available for dystonia and symptomatic treatments in the form of chemodenervation and oral medications provide limited efficacy (Goldman 2015). Mechanistic research studies to identify new targets for long lasting therapy for dystonia are much needed.

WC is an ideal subtype to advance disease mechanism for all forms of focal human dystonia. However, several challenges exist for research studies employing WC dystonia to advance disease mechanism. With an estimated prevalence of 15 per million persons, WC dystonia is a rare disorder that precludes large randomized placebo-controlled design for research studies (Goldman 2015; Ortiz et al. 2018; Nutt et al. 1988; Warner et al. 2000) Another challenge is that the current outcome measures for evaluating efficacy of an intervention consist of examiner-rated scales such as writer’s cramp rating scale (WCRS)(Wissel et al. 1996) and Burke Fahn Marsden (BFM) dystonia and disability scale(Burke et al. 1985). Use of these clinical scales requires specialized practitioners and is susceptible to inter-rater variability. In addition, the coarseness of categorical gradations in these examiner-rated scales may limit the ability to detect a differential response to research manipulations. Lastly, while patient self-reported scales are important for measuring quality of life outcomes, they are not suitable as primary outcome measures in an investigational research study because they are highly susceptible to placebo effect.

Automated motor performance measures offer an opportunity to bypass these limitations. Previous evaluations of writing kinematics using a graphic tablet have reported several writing measures that can distinguish WC from healthy volunteers (HV): mean duration of writing, mean axial pressure, mean stroke frequency, coefficient of variation of peak velocity, and normalized jerk (Zeuner et al. 2007; Siebner et al. 1999; Teresa Jacobson Kimberley et al. 2015; Bradnam et al. 2015; Hermsdörfer et al. 2011; Schenk et al. 2004; Teresa J. Kimberley et al. 2015). Two of these studies additionally compared these kinematic measures to examiner WCRS and patient Arms Dystonia & Disability Scale (Zeuner et al. 2007; Hermsdörfer et al. 2011). The automated measures were found not to correlate with examiner or patient scales; a finding the authors attributed to differing aspects of motor impairments captured by the writing kinematics and rating scales (Zeuner et al. 2007; Hermsdörfer et al. 2011). The inability to correlate automated measures to known examiner and patient scales, however, makes it difficult to apply the research findings to a broader understanding of the clinical disease. Move over, no study to date has evaluated the sensitivity and specificity of these outcome measures to differentiate WC from HV and whether such measures would have sufficient signal to noise ratio to test a research hypothesis with a realistic sample size in this rare disease.

Writing kinematics have also been studied in Parkinson’s disease (PD) patients, with a better understanding of the link between writing measures and clinical disease. Teuling and colleagues used both actual writing samples and simulated writing samples from PD patients to demonstrate a link between the abnormal writing pattern of PD patients and their basal ganglia dysfunction. Specifically, they showed that PD patients show greater variability in the practiced skill of writing compared to HV due to reduced coordination and motor control of finger, wrist and arm movements. The reduced coordination and motor control in the study was shown to result from decreased dynamic modulation of the central movement generator in the basal ganglia which progressively declines with loss of dopamine signal in PD patients (Teulings et al. 1997a). Teulings and colleagues then developed kinematic measures of writing to capture the writing variability of PD patients in a now commercially available software called MovAlyzer. They also showed that a writing measure called normalized jerk could specifically link the clinical manifestation of PD to basal ganglia dysfunction. Given the key role of basal ganglia in the pathophysiology of dystonia, it remains to be seen if the same kinematic measure can also capture clinical manifestations of WC dystonia and thereby link the disease to an objective behavioral measure.

The goal of this study was to identify writing measures suitable for research studies in the WC patients. We focused on three study design features. First, we tested a broad range of automated writing measures generated by the kinematic software MovAlyzer to identify measures that could distinguish WC from HV. Since writing illegibility is a major complaint of individuals with WC (Goldman 2015), we also developed an automated measure of writing legibility using a hand recognition software and evaluated its ability to distinguish WC from HV. The writing legibility measure thus creates an automated but clinically meaningful outcome for patients. Second, we measured the diagnostic performance of the automated writing measures to distinguish WC from HV and its correlation with established dystonia examiner and patient disability rating scales. Third, we evaluated the sample size needed to power a research study using these measures in two different clinical research designs.

## MATERIALS AND METHODS

### Participants

The research study was approved by the Duke University Institutional Review Board. All isolated WC patients with dystonia affecting the right hand were considered eligible for the research study. All enrolled participants were more than 3 months from last botulinum toxin injections and not on oral trihexyphenidyl, a symptomatic treatment for dystonia. Patients were referred from Duke University Movement Disorders Center where they were diagnosed by a Movement Disorder specialist by a standard medical history and neurological examination. All participants provided written consent prior to participation in the research study. All 43 subjects in the study were right-hand dominant with the exception of 1 WC subject (case 7) who reported ambidexterity but wrote with his right hand. Healthy subjects who were right hand-dominant without co-morbid neurological diagnoses and age-matched to a WC patient were recruited from Duke healthy volunteer registry and advertising in the local community.

### Handwriting Measurement Assays

All participants copied the holo-alphabetic sentence “A large fawn jumps quickly over white zinc boxes” 10 times in MovAlyzer kinematic software (Version 6, Neuroscript LLC, Temp, Az) and then 4 times in OneNote text recognition software (Version 2016, Microsoft Office 365). Participants wrote the sentence at self-selected pace and style (cursive or print). Both sentence copying tasks were performed on a digital tablet with a pressure sensitive-pen (Wacom Co, Ltd, Japan). These two assays were performed twice - an initial assessment (“pre-assessment”) and following a 20-minute prolonged writing task (“post-assessment”). During the prolonged writing assessment, subjects performed a writing task in the MovAlyzer software consisting of copying a diverse holo-alphabetic paragraph for 20 minutes.

### Examiner and patient rating scales

All participants were video recorded from the neck down (focused on their right arm). Movement Disorder specialists (B.S. and P.T.) rated the videos using the WCRS (Wissel et al. 1996) while blinded to the identity of the subjects. Examiners also rated all participants’ right arm dystonia severity on the BFM dystonia and disability scale (Burke et al. 1985). Examiners were provided literature on the two rating scales to establish concordant rating. The inter-rater reliability score for the 2 examiners was 0.10 ± 0.04, with WCRS writing tremor (0.43±0.15) and WCRS writing speed (0.49±0.09) subscore measures showing higher inter-rater agreement. All participants self-rated their writing disability on handwriting disability question of BFM Dystonia and Disability scale. The handwriting disability ranges from 0-4 with 0 being normal writing and 4 being unable to grasp to maintain hold on pen. Participants also provided a baseline pain score in their right hand on a 0 to 10 scale with 0 being no pain and 10 being the worst pain.

### Data analysis and statistics

All kinematic variables for each sentence were automatically calculated within the MovAlyzer software and exported for statistical analysis. Each sentence was treated as a single technical replicate (“stroke”). Duration of writing is defined as the time it takes to write a single stroke. The number of peak acceleration points is defined as the number of acceleration peaks both up-going and down-going in a single stroke. Normalized jerk is the change in acceleration during a single stroke normalized for stroke duration and size while normalized y-jerk is the change in acceleration in y-coordinates in a single stroke. Straightness error represents the angular deviation in a single stroke and calculated by taking the average distance to the minimum relative mean square line for each stroke. Writing samples obtained using OneNote software were converted to text using the automated software feature “writing-to-text” conversion. The percent of words correctly translated from writing to text out of 36 total words were scored by an analyst blinded to the group identity.

For statistical analyses, all automated measurements were tested for statistical differences between WC and HV using two-tailed t-tests. All data was initially tested for normality. For data that passed normality, a student t-test was used for two-tailed t-test. For data that failed normality, Mann-Whitney Rank Sum (MWRS) test was used. Correlation analysis between the automated measures and rating scale measures were assessed by Pearson’s correlation coefficient. The correlation coefficient (R) and significance (p-value) reported. For t-test and pearson’s correlation, a p-value <0.05 was considered statistically significant. JMP software (SAS Co, SAS Institute Inc., Cary, NC) was used for these analyses. To characterize the diagnostic performance of automated measures, six logistic regression models were run modelling disease status – one for each automated outcome measure (mean duration, mean peak acceleration points, mean normalized jerk, mean normalized y jerk, mean straightness error, and mean writing legibility). All models were controlled for age and gender. The model formula used for each of 6 automated measures was:

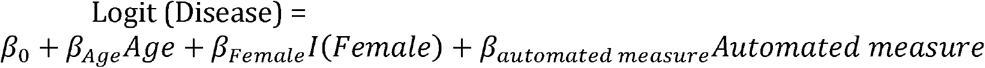

The associated receiver operating curve (ROC) was prepared and plotted for each of the six models. The area under the curve (AUC) was calculated for each of the 6 models. The diagnostic performance analysis was conducted using SAS 9.4 (SAS Institute Inc., Cary, NC). Continuous variables in the text are presented as means (standard deviation [SD]) in the tables and in graph as box plots with medians (25th, 75th percentiles), min and max overlayed on individual subject means Categorical variables such as examiner rating scales are summarized as stacked graphs with number of subjects per score.

### Sample size calculation

Two research designs were used to calculate sample size: 1) A parallel research design to compare differences between HV and WC using a two-tailed unpaired t-test; 2) A single arm cross-over research design was used to detect the intra-subject difference between the two time points (Post-assessment and Pre-assessment) in WC subjects and calculate the standard deviation in a two-tailed paired t-test. In both research designs, an alpha of 0.05 and power of 0.80 was used to estimate the sample size.

## RESULTS

### WC subjects are clinically heterogeneous

21 WC and 22 age-matched HV in equal proportion gender were enrolled in the study with no significant differences in mean age (WC: 59.4(12.2) vs HV: 59.8 (11.6) years, p=0.91, student t-test) or gender (WC: 12M/9F; HV 15M/7F, p=0.45, Chi-square test). WC subjects showed a mean disease duration of 20.4 (12.3) years with range of 4-47 years. 28.6% WC subjects reported simple dystonia while remaining 71.4% endorsed dystonic symptoms with other fine motor tasks in addition to writing (Table 1). None of the healthy subjects reported any dystonic symptoms. With the exception of gender which showed a greater male to female ratio, WC subjects enrolled in this study are representative of the disease population as previously reported (Ortiz et al. 2018; Warner et al. 2000; Nutt et al. 1988).

**Table 1:** Clinical Data of Writer’s Cramp Patients.

| **Subject** | **Gender** | **WC Type** | **Symptom Duration (years)** |
| --- | --- | --- | --- |
| 1 | Male | Simple | 4 |
| 2 | Female | Simple | 10 |
| 3 | Male | Simple | 11 |
| 4 | Male | Simple | 11 |
| 5 | Female | Simple | 20 |
| 6 | Male | Simple | 47 |
| 7 | Male | Dystonic | 6 |
| 8 | Male | Dystonic | 7 |
| 9 | Female | Dystonic | 35 |
| 10 | Female | Dystonic | 12 |
| 11 | Male | Dystonic | 13 |
| 12 | Female | Dystonic | 14 |
| 13 | Female | Dystonic | 16 |
| 14 | Male | Dystonic | 17 |
| 15 | Female | Dystonic | 20 |
| 16 | Male | Dystonic | 22 |
| 17 | Female | Dystonic | 24 |
| 18 | Male | Dystonic | 30 |
| 19 | Male | Dystonic | 30 |
| 20 | Female | Dystonic | 36 |
| 21 | Male | Dystonic | 43 |

### Six automated writing measures can distinguish WC from HV

22 automated measures categorized into 9 kinematic features of writing were analyzed across 43 participants using MovAlyzer software (Table 2). 5 automated measures showed significant difference between WC and HV: duration of writing, number of peak acceleration points, straightness error, normalized y-jerk, and normalized jerk. Interestingly, additional automated measures previously reported in the literature (mean axial pen pressure, average velocity, stroke length, or CV of peak vertical velocity) did not show group level difference. We next assessed writing legibility using an automated handwriting recognition software (Fig. 1A). Using this directly translatable measure of the ability to communicate via handwriting, we found that significantly fewer words were correctly recognized in writing samples from WC subjects compared to HV (Fig. 1B). No differences due to gender were identified across the 6 automated measurements (Suppl. Table 1).

**Table 2:** A comparison of 22 kinematic measures of writing in Healthy and Writer’s Cramp patients.

| Categories | Measures | HV<br>n=22 | WC<br>n=21 | p-value |
| --- | --- | --- | --- | --- |
| **Time of Writing** | Start Time | 0.08(0.09) | 0.12(0.22) | 0.188 |
|  | Duration | 45.80(9.34) | 60.64(28.78) | 0.016* |
|  | Relative Pen Down Duration | 0.66(0.11) | 0.63(0.17) | 0.398 |
| **Velocity of Writing** | Peak Vertical Velocity | -0.06(15.75) | 0.44(20.09) | 0.895 |
|  | Relative Time to Peak Vertical Velocity | 0.36(0.29) | 0.45(0.27) | 0.089 |
|  | Average Absolute Velocity | 3.18(1.59) | 3.21(1.62) | 0.906 |
| **Acceleration of Writing** | Peak Vertical Acceleration | -28.13<br>(513.22) | -69.75<br>(774.52) | 0.738 |
|  | Number of Peak Acceleration Points | 311.01<br>(59.67) | 442.42<br>(208.65) | 0.004** |
| **Position of Writing** | Start Vertical Position | 13.42(4.84) | 15.24(5.41) | 0.057 |
|  | Start Horizontal Position | 4.29(2.76) | 4.52(3.15) | 0.944 |
| **Size of Writing** | Vertical Size | 0.03(1.36) | -0.01(2.66) | 0.930 |
|  | Horizontal Size | 27.74(13.06) | 27.33(13.29) | 0.906 |
|  | Absolute Size | 27.82(12.96) | 27.50(13.21) | 0.906 |
| **Writing Style** | Roadlength | 139.19<br>(60.25) | 177.93<br>(109.69) | 0.196 |
| **Angle of Writing** | Straightness Error | 0.02(0.01) | 0.03(0.05) | 0.006** |
|  | Slant | 0.05(0.20) | 0.00(0.32) | 0.240 |
|  | Relative Initial Slant | 0.54(1.02) | 0.34(1.06) | 0.186 |
| **Dysfluency of Writing** | Normalized Y-Jerk | 4.55×10<sup>6</sup><br>(3.95×10<sup>6</sup>) | 1.87×10<sup>7</sup><br>(3.60×10<sup>7</sup>) | 0.010* |
|  | Normalized Jerk | 1.22×10<sup>7</sup><br>(1.48×10<sup>7</sup>) | 5.15×10<sup>7</sup><br>(9.01×10<sup>7</sup>) | 0.004** |
| **Pressure of Writing** | Average Pen Pressure | 362.77 | 375.39 | 0.792 |

Values are expressed as mean(SD). Group comparison was performed using two-sided unpaired t test or Mann-Whitney rank sum test.\*p<0.05, \*\*p<0.001.

**Figure 1:**
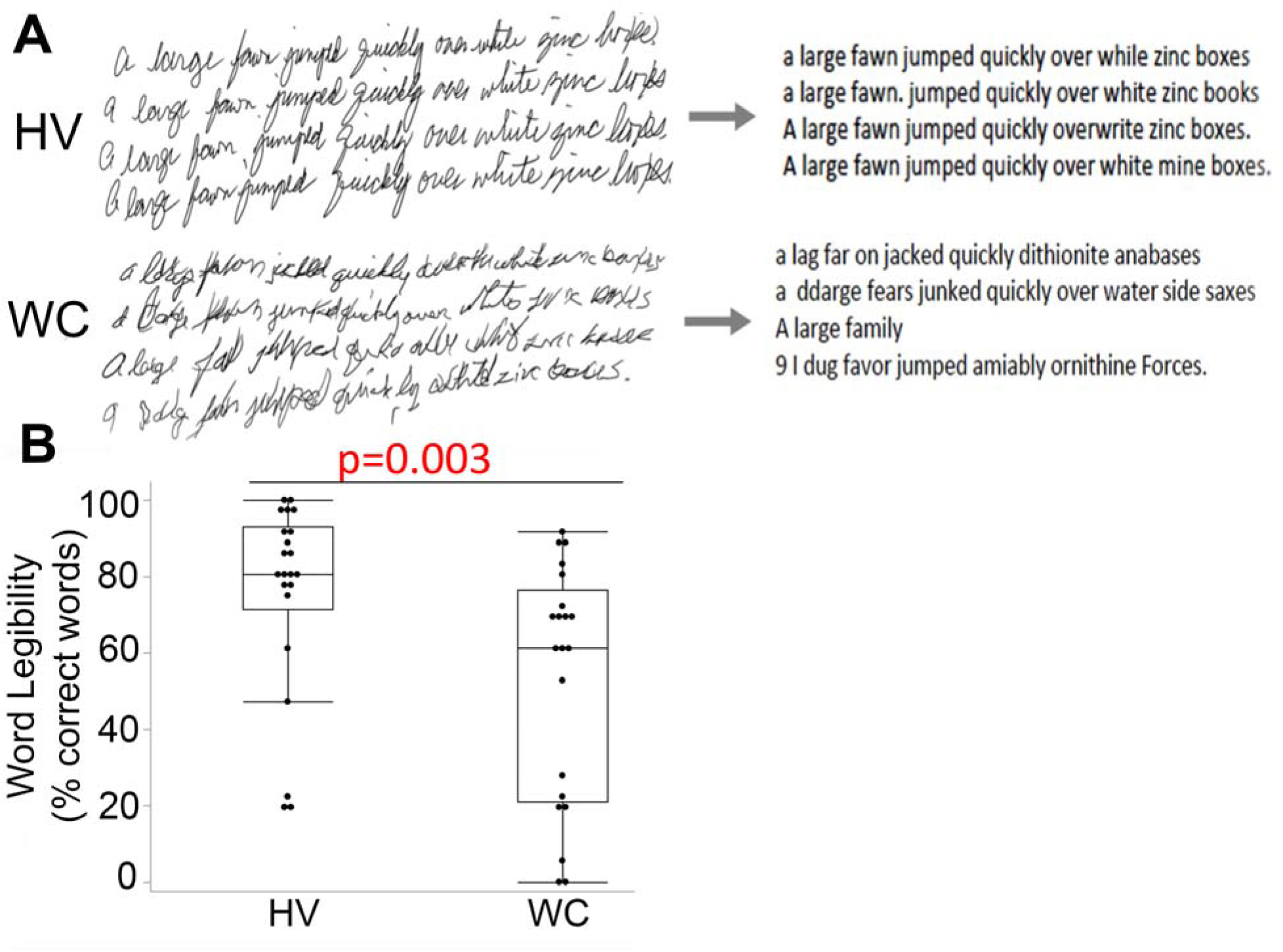
An automated measure of writing legibility can distinguish writer’s cramp (WC) from healthy volunteers (HV). **A)** Representative writing samples from HV and WC patients (left column). An automated writing to text conversion software was used to convert hand-written sentences into text format (right column). B) The percent of correct words in 4 sentences was quantified and expressed as word legibility and expressed as word legibility. P-value from Mann-Whitney rank sum test.

### Six automated measures show high diagnostic performance

We next evaluated the sensitivity and specificity of these 6 measures to distinguish WC from HV using a logistic regression model controlled for age and gender (Fig. 2). The ROC curves shows that all six measures have high sensitivity and specificity to differentiate between the two groups, albeit normalized jerk, normalized y-jerk and writing legibility show higher diagnostic performance (AUC: 0.792-0.775) compared to remaining 3 measures of straightness error, duration and peak acceleration (AUC: 0.720-0.716).

**Figure 2:**
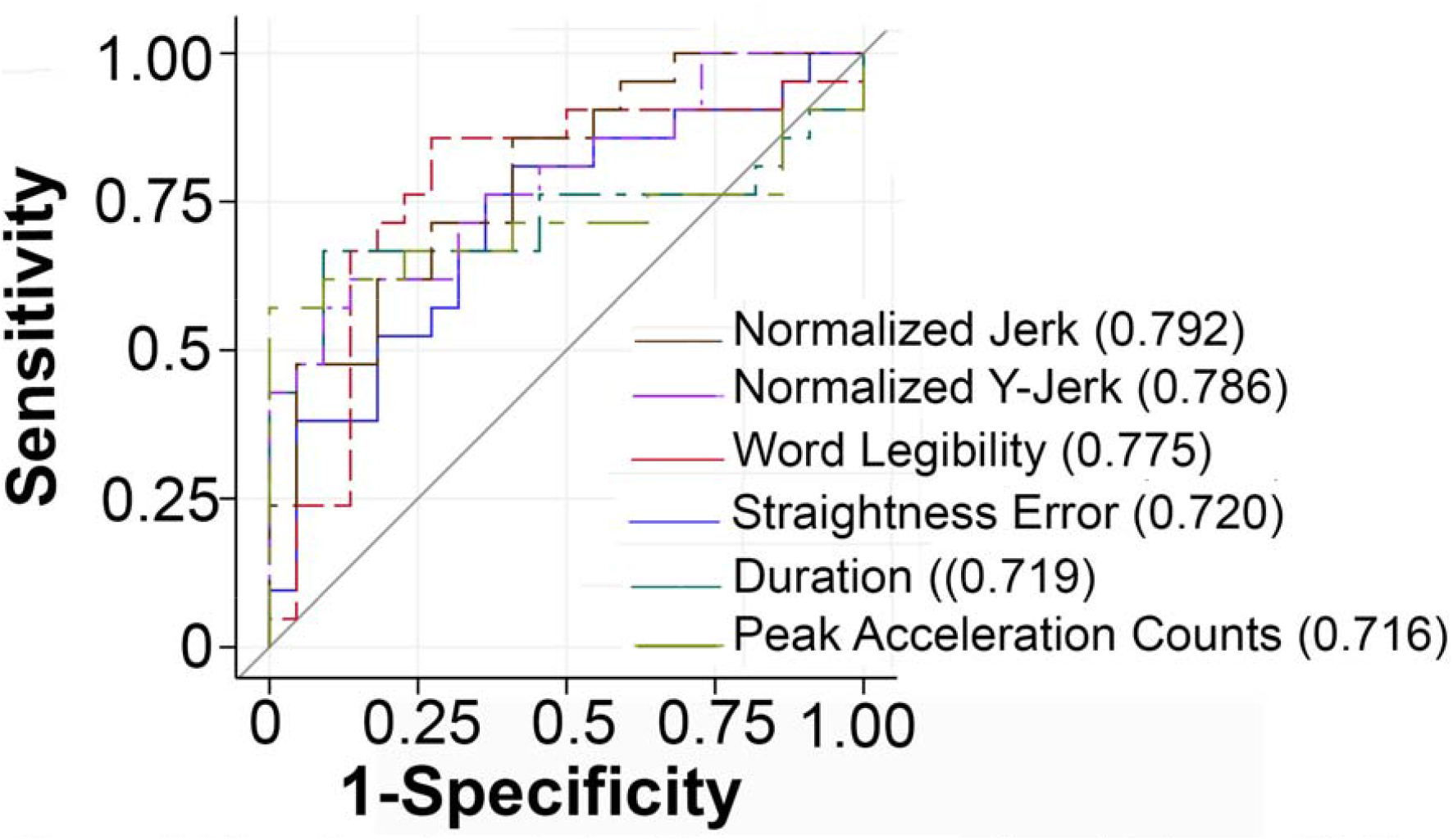
The six automated writing measures show high sensitivity and specificity to predict dystonia status. For each automated measure, a logistic regression test was run to model dystonia status and the diagnostic performance of each measure plotted (ROC curve). ROC curves above the diagonal gray line show high sensitivity and specificity to predict dystonia status. An aggregate measure of diagnostic performance (AUC) is also provided in the parenthesis.

### Automated measures correlate with subscore examiner and patient rating scales of dystonia

Both examiner rating scales (WCRS and BFM right arm dystonia) demonstrate significantly higher measures in WC compared to HV (Suppl. Fig 1). A correlation analysis of the composite scores from the two examiner rating scales with the 6 automated measures did not show significant correlations (Suppl. Table 2) similar to prior studies (Zeuner et al. 2007; Hermsdörfer et al. 2011). However, unlike prior publications, we did find correlations between automated measures and subscore measures of the examiner rating scales in WC patients. Specifically, WCRS part A-writing tremor inversely correlates with writing legibility in WC patients. WCRS part B-writing speed correlates with 5 automated writing measures of duration, number of peak acceleration points, normalized y-jerk and writing legibility in WC patients and normalized jerk in all patients combined (Table 3). The automated measure straightness error shows significant correlation with WC patient’s pain score. BFM writing legibility also inversely correlates with writing legibility measure. The examiner and patient subscore measures failed to show correlation with automated measures in HV.

**Table 3:**
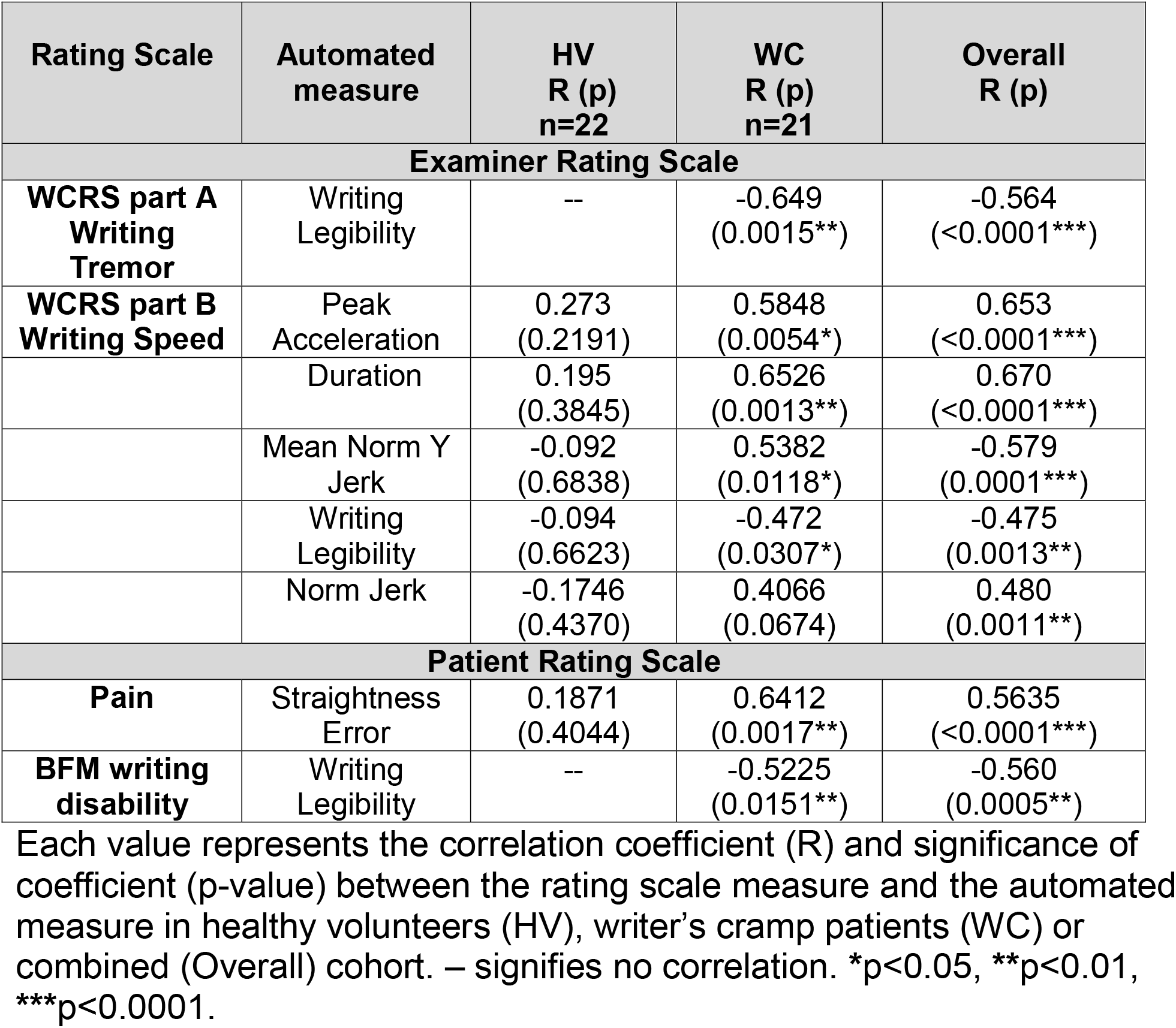
Correlation of automated writing measures with examiner and patient rating scales.

| Rating Scale | Automated measure | HV<br>R (p)<br>n=22 | WC<br>R (p)<br>n=21 | Overall<br>R (p) |
| --- | --- | --- | --- | --- |
| **Examiner Rating Scale** |  |  |  |  |
| **WCRS part A**<br>**Writing**<br>**Tremor** | Writing Legibility | -- | -0.649<br>(0.0015**) | -0.564<br>(<0.0001***) |
| **WCRS part B**<br>**Writing Speed** | Peak Acceleration | 0.273<br>(0.2191) | 0.5848<br>(0.0054*) | 0.653<br>(<0.0001***) |
|  | Duration | 0.195<br>(0.3845) | 0.6526<br>(0.0013**) | 0.670<br>(<0.0001***) |
|  | Mean Norm Y Jerk | -0.092<br>(0.6838) | 0.5382<br>(0.0118*) | -0.579<br>(0.0001***) |
|  | Writing Legibility | -0.094<br>(0.6623) | -0.472<br>(0.0307*) | -0.475<br>(0.0013**) |
|  | Norm Jerk | -0.1746<br>(0.4370) | 0.4066<br>(0.0674) | 0.480<br>(0.0011**) |
| **Patient Rating Scale** |  |  |  |  |
| **Pain** | Straightness Error | 0.1871<br>(0.4044) | 0.6412<br>(0.0017**) | 0.5635<br>(<0.0001***) |
| **BFM writing disability** | Writing Legibility | -- | -0.5225<br>(0.0151**) | -0.560<br>(0.0005**) |

Each value represents the correlation coefficient (R) and significance of coefficient (p-value) between the rating scale measure and the automated measure in healthy volunteers (HV), writer's cramp patients (WC) or combined (Overall) cohort. – signifies no correlation. \*p<0.05, \*\*p<0.01, \*\*\*p<0.0001.

### Sample size needed to detect group differences varies widely across the 6 automated measures

We next calculated the sample size needed for each of the 6 automated measures to detect a clinical effect in 2 common clinical research designs. A single arm cross-over research design in WC patients would allow for study of the effect of a research manipulation or clinical intervention in the same disease patients. Although definition of a clinically meaningful effect size can vary widely as a consequence of study aims, we used a medium Cohen’s d-effect size of 0.3 to evaluate sample size in this research design. The estimated sample size of WC patients needed to see a clinical effect for each measure was: writing legibility-32; duration-57; peak acceleration-58; normalized jerk-84; normalized y-jerk – 123; and straightness error-300. A parallel research design in WC and HV would compare the effect of a research manipulation or clinical intervention between the two groups. The estimated sample size of each group needed to see a clinical effect for each measure is: writing legibility-23; peak acceleration-18; duration-26; normalized jerk-25; normalized y-jerk – 30; and straightness error-43. Overall, the sample size calculations across the 2 research designs show a wide range of sample sizes with smaller sample sizes estimated for measures of writing legibility, duration and peak acceleration and larger sample sizes estimated for measures of normalized jerk, normalized y-jerk and straightness error. Furthermore, of the 2 research designs, a parallel research design demonstrates lower sample size requirements to see clinical effects across all automated measures.

## DISCUSSION

In summary, we find that three objective, automated writing measures (writing legibility, duration of writing, and number of peak accelerations points) that can significantly differentiate WC subjects from HV, correlate with clinical features of dystonia recognized by expert examiners using validated scales and achieve a realistic sample size for use in a clinical research study.

To our knowledge, our study is the first to evaluate the utility of a consumer accessible writing recognition software as an outcome measure for writing legibility in WC. Previous studies have used circle drawing to evaluate the accuracy of writing movements and have shown group level differences (Murase et al. 2005; Hermsdörfer et al. 2011; Zeuner et al. 2007). The automated writing legibility measure in this study captures group-wise differences in writing accuracy previously reported (Zeuner et al. 2007; Murase et al. 2005; Hermsdörfer et al. 2011), but is a more proximal measure of the clinically meaningful outcome of improving writing legibility. Furthermore, of the 3 automated measures identified here, writing legibility requires the smallest estimated sample size to detect a moderate effect size in a research study, thus making it a realistic outcome measure in this patient population.

Using kinematic measures of writing, WC subjects also exhibit significantly greater dysfluent events compared to HV. Similar to PD study (Teulings et al. 1997b), our study also found normalized jerk measure to distinguish WC from HV. However, we found that the measure peak accelerations captured a similar phenomenology but the sample size needed in a research study will be more achievable in this patient population. Our study also found that WC patients take significantly more time to write than HV as supported by the automated measurement duration. The increased duration finding is consistent with a previously published study which found that writing duration distinguished WC from HV in a repeated sentence copying assessment (Hermsdörfer et al. 2011). Correlational analyses also demonstrate that writing duration captures the same feature as evaluated by the traditional examiner-rated writing speed measure. As expected, the correlation between examiner-rated dystonia scales and automated measure were specific to WC patients with no correlations seen in HV. The correlation between straightness error and patient pain suggests that straightness error may be capturing secondary compensation and pain resulting from their underlying dystonia rather than the primary movement itself. The inverse correlation between patient reported BFM writing disability and automated writing legibility measure further supports that the automated measure captures a meaningful clinical outcome for patients. While our findings reinforce some prior observations, our study did not identify group differences in other primary measures that have previously been reported such as mean stroke frequency, mean axial pressure, and CV of peak velocity (Siebner et al. 1999; Hermsdörfer et al. 2011; Teresa J. Kimberley et al. 2015; Zeuner et al. 2007). However, we note that there are methodological differences between our study and these studies. Overall, our data supports that the 6 automated writing measures show high sensitivity and specificity to distinguish WC from HV and correlate with features of dystonia and disability captured by traditional examiner and patient rating scales.

Future studies should evaluate if the automated writing measures may also reflect dystonia pathophysiology. A link between kinematic measures of writing and basal ganglia dysfunction in PD patients was previously demonstrated. It would be worthwhile to determine if changes in basal ganglia function in WC dystonia as well as other form of focal dystonia correlate with the automated writing measures identified in this study.

In conclusion, we have identified three automated writing measures: a novel writing legibility measure and kinematic measures of duration of writing and peak acceleration points not widely used in WC dystonia research studies. At present, therapeutic opportunities for individuals affected by WC are limited and affected individuals suffer from both an impaired ability to communicate via writing as well as pain. A critical step towards improving the therapeutic landscape for WC and broadly other forms of focal dystonias are validated outcome measures suitable for testing small populations and relevant to clinically meaningful outcomes. It is our hope that use of validated automated outcome measures such as the ones presented in this research study can allow for more effective research design to advance disease mechanism and therapies in this disabling and rare disorder.

## Data Availability

Data is available upon request from the primary author.

## AUTHOR CONTRIBUTIONS

NBP was involved in all aspects of the research project from conceptualizing the study, to data collection, statistical analysis and manuscript writing. AM, HAK, and JW were involved in designing and performing the statistical analysis of the data. BS and PT served as blinded examiners in the study and reviewed and critiqued the manuscript. LA was involved in the data analysis and reviewed and critiqued the manuscript. ML was involved in designing the statistical analysis as well as reviewed and critiqued the manuscript. NC conceptualized the research study, reviewed and critiqued the data analysis and assisted with manuscript writing.

## ACKNOWLEDGEMENTS

The authors would like to thank Amber Holden, Ashley Pifer and Kelsey Ling who served as Clinical Research Coordinators for this study.

## REFERENCES

Albanese, Alberto, Kailash Bhatia, Susan B. Bressman, Mahlon R. Delong, Stanley Fahn, Victor S.C. Fung, Mark Hallett, et al. 2013. “Phenomenology and Classification of Dystonia: A Consensus Update.” Movement Disorders 28 (7): 863–73. https://doi.org/10.1002/mds.25475.

Bradnam, Lynley V, Lynton J Graetz, Michelle N Mcdonnell, and C Michael. 2015. “Anodal Transcranial Direct Current Stimulation to the Cerebellum Improves Handwriting and Cyclic Drawing Kinematics in Focal Hand Dystonia” 9 (May): 1–9. https://doi.org/10.3389/fnhum.2015.00286.

Burke, Robert E., Stanley Fahn, C. David Marsden, Susan B. Bressman, Carol Moskowitz, and Joseph Friedman. 1985. “Validity and Reliability of a Rating Scale for the Primary Torsion Dystonias.” Neurology 35 (1): 73–77.

Goldman, Jennifer G. 2015. “Writer’s Cramp.” Toxicon 107: 98–104. https://doi.org/10.1016/j.toxicon.2015.09.024.

Hallett, Mark. 2006. “Pathophysiology of Writer’s Cramp.” Human Movement Science 25 (4–5): 454–63. https://doi.org/10.1016/j.humov.2006.05.004.

Hermsdörfer, Joachim, Christian Marquardt, Alexandra S. Schneider, Waltraud Fürholzer, and Barbara Baur. 2011. “Significance of Finger Forces and Kinematics during Handwriting in Writer’s Cramp.” Human Movement Science 30 (4): 807–17. https://doi.org/10.1016/j.humov.2010.04.004.

Jedynak, Pierre C., Christine Tranchant, and Diederik Zegers de Beyl. 2001. “Prospective Clinical Study of Writer’s Cramp.” Movement Disorders 16 (3): 494–99. https://doi.org/10.1002/mds.1094.

Kimberley, Teresa J., Rebekah L. S. Schmidt, Mo Chen, Dennis D. Dykstra, and Cathrin M. Buetefisch. 2015. “Mixed Effectiveness of RTMS and Retraining in the Treatment of Focal Hand Dystonia.” Frontiers in Human Neuroscience 9 (July): 1–11. https://doi.org/10.3389/fnhum.2015.00385.

Kimberley, Teresa Jacobson, Michael R. Borich, Rebekah L. Schmidt, James R. Carey, and Bernadette Gillick. 2015. “Focal Hand Dystonia: Individualized Intervention with Repeated Application of Repetitive Transcranial Magnetic Stimulation.” Archives of Physical Medicine and Rehabilitation 96 (4). https://doi.org/10.1016/j.apmr.2014.07.426.

Murase, Nagako, John C Rothwell, Ryuji Kaji, Ryo Urushihara, Kazumi Nakamura, Nobuki Murayama, Tomohiko Igasaki, Miyuki Sakata-igasaki, Tatuya Mima, and Akio Ikeda. 2005. “Subthreshold Low-Frequency Repetitive Transcranial Magnetic Stimulation over the Premotor Cortex Modulates Writer ‘ s Cramp” 128 (1): 104–15. https://doi.org/10.1093/brain/awh315.

Nutt, John G., Manfred D. Muenter, Arnold Aronson, Leonard T. Kurland, and L. Joseph Melton. 1988. “Epidemiology of Focal and Generalized Dystonia in Rochester, Minnesota.” Movement Disorders. https://doi.org/10.1002/mds.870030302.

Ortiz, Rebekka, Filip Scheperjans, Tuomas Mertsalmi, and Eero Pekkonen. 2018. “The Prevalence of Adult-Onset Isolated Dystonia in Finland 2007-2016.” PLoS ONE. https://doi.org/10.1371/journal.pone.0207729.

Schenk, Thomas, Barbara Bauer, Birgit Steidle, and Christian Marquardt. 2004. “Does Training Improve Writer’s Cramp? An Evaluation of a Behavioral Treatment Approach Using Kinematic Analysis.” Journal of Hand Therapy 17 (3): 349–63. https://doi.org/10.1197/j.jht.2004.04.005.

Siebner, Hartwig Roman, J. M. Tormos, A. O. Ceballos-Baumann, C. Auer, M. D. Catala, B. Conrad, and A. Pascual-Leone. 1999. “Low-Frequency Repetitive Transcranial Magnetic Stimulation of the Motor Cortex in Writer’s Cramp.” Neurology 52 (3): 529–37.

Teulings, Hans Leo, JoséL. Contreras-Vidal, George E. Stelmach, and Charles H. Adler. 1997a. “Parkinsonism Reduces Coordination of Fingers, Wrist, and Arm in Fine Motor Control.” Experimental Neurology. https://doi.org/10.1006/exnr.1997.6507.

———. 1997b. “Parkinsonism Reduces Coordination of Fingers, Wrist, and Arm in Fine Motor Control.” Experimental Neurology 146 (1): 159–70. https://doi.org/10.1006/exnr.1997.6507.

Warner, T., L. Camfield, C. D. Marsden, A. H. Nemeth, N. Hyman, D. Harley, J. Wissel, et al. 2000. “A Prevalence Study of Primary Dystonia in Eight European Countries.” Journal of Neurology. https://doi.org/10.1007/s004150070094.

Wissel, J, C Kabus, R Wenzel, S Klepsch, U Schwarz, A Nebe, L Schelosky, and U Scholz. 1996. “Botulinum Toxin in Writer ‘ s CramplJ: Objective Response Evaluation in 31 Patients.” Journal of Neurology 61: 172–75.

Zeuner, Kirsten E., Martin Peller, Arne Knutzen, Iris Holler,Alexander Münchau, Mark Hallett, Günther Deuschl, and Hartwig R. Siebner. 2007. “How to Assess Motor Impairment in Writer’s Cramp.” Movement Disorders 22 (8): 1102–9. https://doi.org/10.1002/mds.21294.

