## Supplementary Material for "Designing research studies in writer’s cramp dystonia: an analysis of automated writing measures"

**SUPPLEMENTARY METHODS:**

**Examiner Rating Scales:** The WCRS has two relevant subscales with part A scoring writing movement (posture, latency, tremor, 28 maximum points) and part B scoring writing speed (2 maximum points). Categorical values from 0 to 2 are assigned with 0 being normal and 2 being marked impairment. The BFM scale assesses the severity and nature of provocation for dystonia in all body parts. Since raters were provided video of only the right arm during writing, only the BFM right arm dystonia severity subscore was used. This BFM arm dystonia scale assigns values from 0 to 4, 0 being normal and 4 being severe, no useful grasp.

**SUPPLEMENTARY DATA:**

**Supplementary Table 1: 6 Automated Writing measures by gender**

| **Group** | **HV** | **HV** |  | **WC** | **WC** |  |
| --- | --- | --- | --- | --- | --- | --- |
| **Gender** | **Male**  **(n=15)** | **Female**  **(n=7)** | **p-value** | **Male**  **(n=12)** | **Female**  **(n=9)** | **p-value** |
| Norm Jerk | 8.57x10^7^  (9.28x10^6^) | 2.29x10^7^  (1.76x10^7^) | 0..097 | 5.94x10^7^  (7.51x10^7^) | 4.11x10^7^  (5.50x10^7^) | 0.594 |
| Norm Y-Jerk | 4.02x10^6^  (2.81x10^6^) | 6.17x10^6^  (2.91x10^6^) | 0.153 | 2.53x10^7^  (3.35x10^7^) | 9.86x10^6^  (8.41x10^6^) | 0.374 |
| Duration | 44.28  (7.69) | 49.27  (9.75) | 0.294 | 63.79  (30.93) | 56.45  (14.98) | 0.483 |
| Peak Acceleration | 312.09  (39.86) | 309.65  (62.23) | 0.931 | 460.95  (206.45) | 417.71  (152.22) | 0.603 |
| Straightness Error | 0.01  (0.00) | 0.02  (0.01) | 0.071 | 0.04  (0.04) | 0.02  (0.01) | 0.594 |
| Writing Legibility | 69.65  (27.69) | 90.74  (8.55) | 0.054 | 44.44  (36.43) | 64.51  (17.45) | 0.114 |

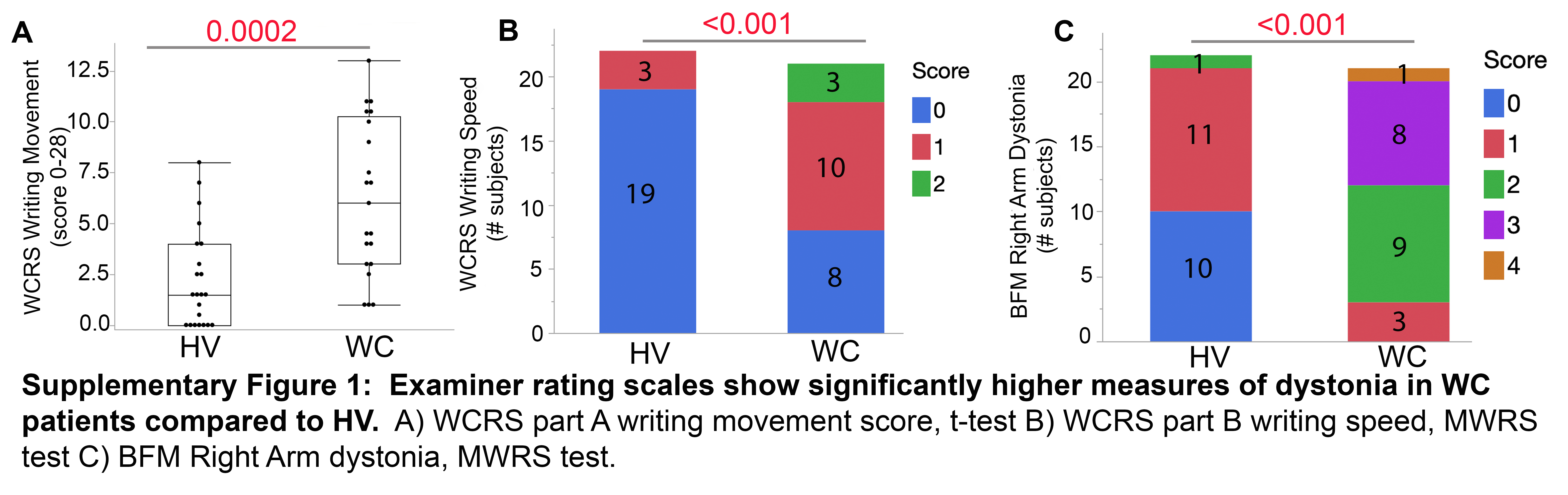

**Supplementary Table 2: Correlation of automated writing measures with examiner rated composite measures**

| **Rating Scale** | **Automated measures** | **HV**  **R (p)**  **n=22** | **WC**  **R (p)**  **n=21** | **Overall**  **R (p)** |
| --- | --- | --- | --- | --- |
| **Examiner Rating Scales** | | | | |
| **WCRS part A Movement score** | Norm Jerk | -0.0973  (0.6665) | -0.116  (0.6154) | 0.129  (0.4098) |
|  | Norm Y-Jerk | 0.0401 (0.8594) | -0.131  (0.5706) | 0.111  (0.4769) |
|  | Duration | -0.0831 (0.7133) | -0.081  (0.7267) | 0.146  (0.3494) |
|  | Peak Acceleration | -0.153 (0.4966) | -0.034 (0.8834) | 0.210  (0.1764) |
|  | Straightness Error | -0.0595  (0.7926) | -0.185 (0.4227) | 0.039  (0.8023) |
|  | Writing Legibility | 0.311  (0.1592) | 0.168  (0.4672) | -0.033  (0.8336) |
| **BFM Right arm dystonia** | Norm Jerk | -0.009 (0.9700) | 0.0946  (0.6835) | 0.348  (0.0223*) |
|  | Norm y-Jerk | 0.046 (0.8391) | 0.220  (0.3382) | 0.385  (0.0108*) |
|  | Duration | 0.128  (0.569) | 0.048  (0.8351) | 0.336  (0.0273*) |
|  | Peak Acceleration | 0.101  (0.6545) | -0.037  (0.8736) | 0.3508  (0.0211*) |
|  | Straightness Error | -0.2240  (0.3162) | 0.319  (0.1590) | 0.362  (0.0172*) |
|  | Writing Legibility | 0.073  (0.7481) | -0.173  (0.4528) | -0.339  (0.0263*) |

Each value represents the correlation coefficient (R) and significance of coefficient (p-value) between the rating scale measure and the automated measure in healthy volunteers (HV), writer’s cramp patients (WC) or combined (Overall) cohort. *****p<0.05, ******p<0.01, *******p<0.0001.
